# Disease signatures in the gut metagenome of a prospective family cohort of inflammatory bowel disease

**DOI:** 10.1101/2023.12.10.23299783

**Authors:** Malte Rühlemann, Silvio Waschina, Eike Matthias Wacker, Philipp Rausch, Ana Schaan, Zaira Zafroon, Katrin Franzpötter, Gunnar Jacobs, David Ellinghaus, Robert Kruse, Daniel Bergemalm, Jonas Halfvarson, Malte Ziemann, Siegfried Görg, Wolfgang Lieb, Stefan Schreiber, Mathilde Poyet, Corinna Bang, Andre Franke, Mathieu Groussin

## Abstract

Inflammatory bowel disease (IBD) is associated with dysbiotic microbiomes. However, whether microbiomes of family members of IBD patients harbour microbial disease signatures of IBD is unknown. Here, we generate shotgun metagenomic data of an IBD family cohort and treatment-naive IBD cases, which we combine with published IBD metagenomes, to perform a meta-analysis of IBD-microbiome associations. Our study reveals microbial shifts that are specific to IBD or IBD subtypes (Crohn’s disease and ulcerative colitis). We find that IBD cohorts share signatures of association with disease irrespective of geography, and we report novel species-level identifications of microbial taxa that are universal markers of Crohn’s disease. We further demonstrate that the microbiome of healthy family members at high risk for IBD represent transitional states between the microbiome of unrelated healthy controls and IBD cases, and harbor diversity, compositional and ecological features that are also observed among IBD microbiomes. Overall, our study provides valuable insights into the intricate relationship between the gut microbiome and IBD, with implications for better predicting disease onset among individuals with high susceptibility for IBD.

## Introduction

Inflammatory Bowel Disease (IBD), encompassing conditions such as Crohn’s disease (CD) and ulcerative colitis (UC), represents a complex and debilitating group of chronic inflammatory disorders affecting the gastrointestinal tract (Alatab et al., 2020; Baumgart & Sandborn, 2007; Lönnfors et al., 2014) The pathogenesis of IBD involves intricate interactions among genetic predisposition, environmental factors, immune system dysregulation, and the gut microbiome (Baumgart & Carding, 2007; Graham & Xavier, 2020; Plichta et al., 2019). Previous microbiome studies identified multiple taxonomic, genomic or metabolic signatures of disease compared to healthy individuals, e.g. with a decrease in *Faecalibacterium* abundance and elevated levels of Enterobacteriacea and *Veillonella* (Häsler et al., 2017; IBDMDB Investigators et al., 2019; Ma et al., 2022; Mallick et al., 2021; Schirmer et al., 2019). Specific bacterial metabolites, such as the short-chain fatty acid butyrate, have also been shown to be altered in IBD and to be associated with clinical remission following anti-TNF therapy (Aden et al., 2019; Meade et al., 2023). The transfer of fecal microbiome from IBD patients into germ-free mice elevates pro-inflammatory T cell responses (*e.g.*, Th17) and exacerbates disease symptoms in a mouse model of colitis (Britton et al., 2019). While caution is advised in interpreting human microbiota transfer experiments into mice (Walter et al., 2020), these findings imply that the microbiome may convey IBD disease phenotypes and play a role in IBD development and progression (Ni et al., 2017).

Despite extensive sequencing studies on the IBD microbiome, reported associations between IBD and specific microbes vary across studies (Gevers et al., 2014; Halfvarson et al., 2017; IBDMDB Investigators et al., 2019; Papa et al., 2012). These inconsistencies arise from differences in sample size, clinical heterogeneity, significant inter-individual and inter-population variations in microbiome compositions, and the construction of cross-sectional case-control studies that limit our ability to account for temporal variations in the microbiome within individuals (Gibbons et al., 2018; Schirmer et al., 2018). These inconsistent findings underscore the need for systematic approaches, such as meta-analyses or the use of validation cohorts, to reveal general patterns of disease-associated microbiome shifts (Abbas-Egbariya et al., 2022; Abdel-Rahman & Morgan, 2023; Duvallet et al., 2017). Recent clinical and methodological efforts greatly improved our understanding of the microbial factors that associate with IBD, incorporating multi-center or meta-analyses of both cross-sectional and longitudinal microbiome datasets covering various intestinal biospecimens and omics profiling (Ma et al., 2022).

The majority of IBD microbiome studies used 16S rRNA amplicon sequencing, limiting our ability to identify microbial disease signatures at fine taxonomic resolutions (e.g., species or strain) due to the restricted information in amplicon data and the incompleteness of 16S-based reference taxonomic databases (Abbas-Egbariya et al., 2022; Ma et al., 2022). For example, unclassified Lachnospiraceae bacteria are often associated with IBD, but the specific species involved remain unknown, hindering follow-up experimental investigations into their functional role in disease (Ma et al., 2022). As of now, the availability of IBD metagenomic datasets for the community is limited in terms of study number, sample size, and sequencing depth (IBDMDB Investigators et al., 2019; Vich Vila et al., 2018). Compared to 16S, metagenomic approaches provide the opportunity to build dataset-specific reference genome collections, profile precise taxonomies, capture functional repertoires (e.g., through gene family or pathway profiling), and identify bacterial genomic markers of host phenotypes down to the resolution of structural variants or single nucleotide polymorphisms (Blanco-Míguez et al., 2023; Olm et al., 2021; Rühlemann et al., 2022). As such, accessing large-scale longitudinal metagenomic datasets from IBD clinical cohorts is crucial to reveal and experimentally validate the contributions of the gut microbiome to IBD etiology and prognosis.

Previous IBD cohorts mainly enrolled patients with active disease, making it challenging to understand the sequential events leading to dysbiotic microbiomes and to discern whether changes in the microbiome are attributable to inflammation, drugs, or comorbidities (Raygoza Garay et al., 2023; Wright et al., 2015). First-degree relatives of IBD patients face an increased 5% lifetime risk of developing IBD (Halme, 2006; Russell & Satsangi, 2008), and exhibit higher levels of intestinal inflammation and impaired intestinal barrier function (Keita et al., 2018; Lee et al., 2019). The extent to which microbiomes contribute to this increased susceptibility and display signatures of early disease development is poorly characterized. Interestingly, previous studies demonstrated positivity for various anti-microbial antibodies, such as ASCA (anti-Saccharomyces cerevisiae antibodies) or ALCA (anti-laminaribioside carbohydrate antibodies) in relatives of CD patients (Michielan et al., 2013; Vermeire et al., 2001). In addition, a recent 16S-based analysis of a prospective cohort of CD cases and their first-degree relatives identified taxonomic markers of future onset of CD (Raygoza Garay et al., 2023). This body of evidence points to an early causative role of the gut microbiome in IBD development.

Here, we generated metagenomic data from hundreds of IBD patients and IBD family members with extensive clinical and environmental metadata, and performed a cross-country meta-analysis to investigate associations between microbiome features and disease. We reveal novel microbiome markers of IBD, including features that delineate cases from controls, or that are specific to either CD, UC or IBD, as well as reduced auxotrophic capacity of the IBD microbiome, which suggest lower metabolic cross-feeding interactions in dysbiotic microbiomes.

## Main text

### Cohort description

The Inflammatory Bowel Disease Family Cohort (IBD-FC) cohort is a German nationwide, prospective cohort study, which was established at Kiel University, Germany, in 2013, recruiting individuals affected by IBD, either CD or UC, together with their IBD-unaffected family members, to build a resource for the pro- and retrospective assessment of (incident) IBD cases. After the initial baseline sampling, participants are invited for follow-up appointments and biosample collection every 2-3 years or, in case of a newly diagnosed IBD case, soon after the initial diagnosis.

We sequenced and analyzed fecal metagenomes of 199 individuals diagnosed for IBD (n_CD_ = 99; n_UC_ = 100) and of 296 in IBD-unaffected family members (family controls, FC) in the IBD-FC cohort. We further generated metagenomes from unrelated healthy population controls (HC; n=344). We also generated metagenomes for 50, 53 and 147 of the CD, UC and FC individuals at a second time point, from ∼ 2 years after the initial sampling was performed. We additionally generated metagenomes from a cohort of treatment-naïve CD- and UC-affected individuals (n_CD=17 and n_UC=16) from Sweden, together with unrelated healthy controls (n=17) and individuals with bowel-complaints not related to IBD (symptomatic controls, SC; n=16). Further, as an additional cohort with longitudinal data, we included a subset of the HMP2 dataset from the USA for which a second sampling timepoint ∼ 180 days after initial sampling was available, resulting in the inclusion of 89 individuals (n_CD_ = 39, n_UC_ = 25, n_HC_ = 25) (IBDMDB Investigators et al., 2019). The overall dataset analyzed in this study includes n = 155 and n = 141 CD- and UC-affected individuals, respectively, in addition to n = 682 IBD-unaffected individuals, totaling samples from n = 978 individuals from three countries.

### IBD and subtypes harbor strong, yet distinct microbial signatures

**Figure 1:**
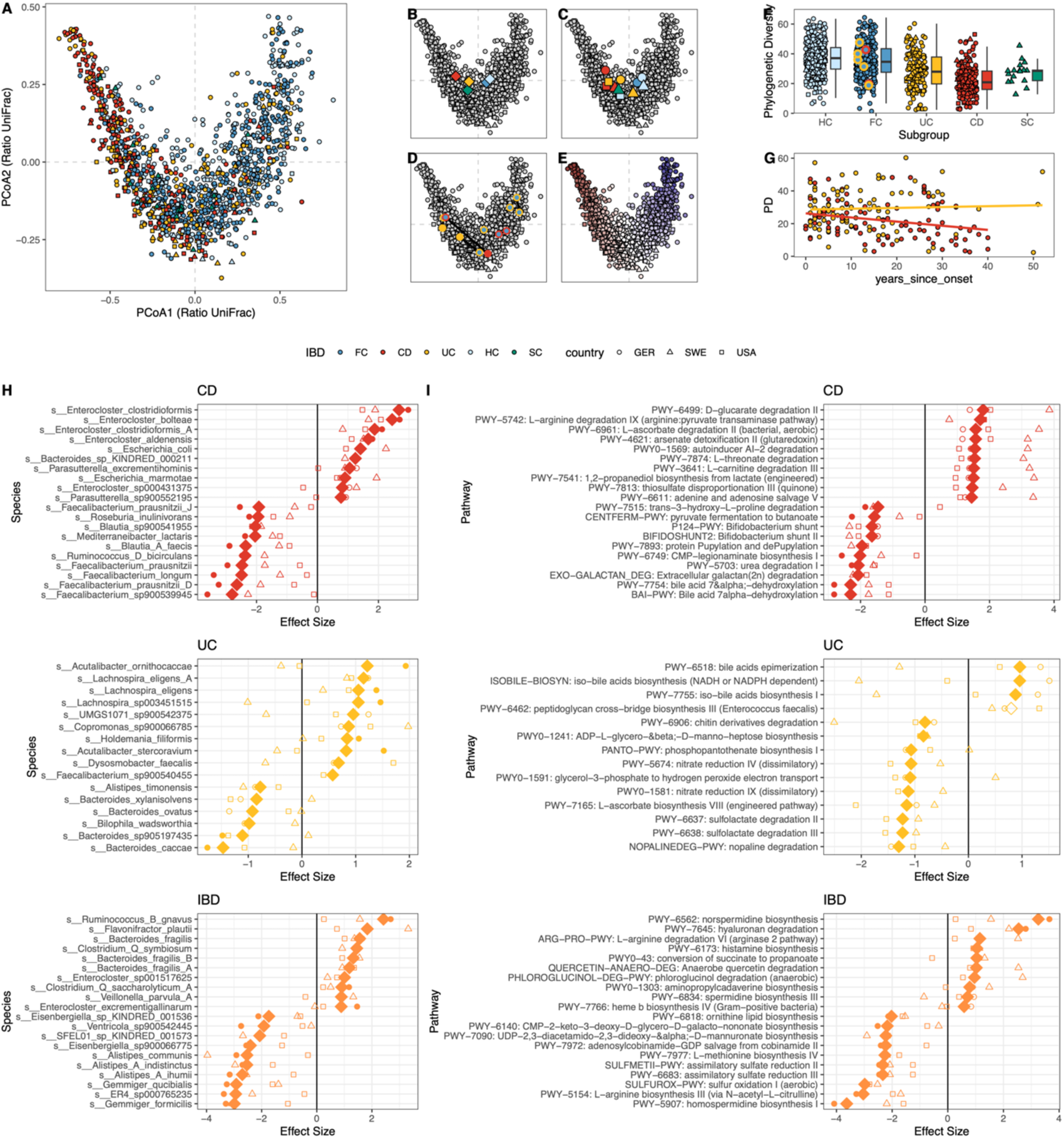
Our meta-analysis of gut metagenomes reveals microbiome changes that are associated with IBD and IBD subtypes. (A) Multivariate analysis (PCoA) plot based on Ratio UniFrac distances. (B) Subgroup centroids; HC = healthy population controls, FC = family controls, CD = Crohn’s disease, UC = Ulcerative colitis, SC = symptomatic controls (SWE). (C) Country-wise subgroup centroids. (D) IBD incident cases in the IBD-FC cohort following baseline sampling/ Samples from multiple timepoints are connected by a line. Blue-filled circles are pre-onset samples, yellow/red filled circles are post-diagnosis. (E) Samples are colored by phylogenetic diversity (high/low diversity: blue/red, respectively). (F) Comparison of phylogenetic diversity across subgroups. (G) Phylogenetic diversity decreases in CD over time since disease onset (P=0.0146), but not in UC (P=0.78); linear regression model. (H) Top 20 bacterial species with the strongest abundance shifts that are specific to either a particular disease subtype (CD, UC) or to general IBD, compared to HC. (I) Top 20 pathways with the strongest abundance shifts that are specific to either a particular disease subtype (CD, UC) or to general IBD, compared to HC. The full list of bacterial and pathway hits can be found in Supplementary Table 2 and Supplementary Figure 3.

We generated a catalog of metagenome-assembled genomes (MAGs) that we clustered into non-redundant species-level genome bins (SGBs), which were taxonomically and functionally annotated (See Methods). This SGB catalog served as a mapping reference for estimating taxonomic and pathway abundance profiles (See Methods). At the aggregate level, we observed a strong shift in microbial compositions among individuals affected by IBD when compared to both FCs and HCs, with the most pronounced differences observed in CD (Figure 1A and B). At the level of individual countries, this shift was consistently reproduced (Fig. 1C). We also observed country-specific signatures of microbial diversities, with USA HC microbial compositions being close to those of groups affected by disease and intestinal symptoms in Sweden and Germany (UC microbiomes). This pattern is also confirmed by a generally lower phylogenetic diversity in USA HC microbiomes. This heterogeneity between populations confirms the importance of performing cross-geography analyses to reveal universal and replicated association signatures of IBD.

Within the FC group, seven individuals were diagnosed with IBD (n_CD_ = 2, n_UC_ = 5) following initial recruitment (Figure 1D). While we include these onset cases in our analyses and highlight them in our results (Figure 1D), the low incidence sample size precluded dedicated statistical analysis – future analyses with additional onset cases will be needed in the future to allow longitudinal microbiome comparisons before and after disease manifestation.

We further found that shifts in microbiome compositions are linked to a reduction in phylogenetic diversity within samples (Figure 1E). This decrease in diversity is further underscored by between-group comparisons, showing reduced diversity in both IBD subtypes (P_CD_ = 1.06 x 10^-29^, P_UC_ = 1.40 x 10^-13^), with no significant difference being observed between HC and FC (Figure 1F). Moreover, substantial differences in phylogenetic diversity were found between HCs from different countries, with the USA HC cohort exhibiting lower phylogenetic diversity compared to their European counterparts (Suppl. Figure S1). This emphasizes the need for proper healthy control cohorts when performing multi-country comparisons of microbiome datasets, which need to account for geographic disparities in the diversity and composition of the baseline healthy microbiome. Interestingly, a decline in phylogenetic diversity over time since disease onset was specifically observed for CD (P = 0.0146, linear regression controlling for sex and BMI; Figure 1G), suggesting a potential association between disease progression and the microbial dysbiosis associated with CD, whereas no such correlation was found for UC (P = 0.78, controlling for sex and BMI; Figure 1G).

We conducted a differential abundance analysis of microbial taxonomies at the species level (Figure 1H) and of microbial pathways (Figure 1I) across all three included cohorts, using a meta-analysis approach that is based on likelihood-based measures (See Methods). This analysis aimed to determine whether observed changes are generally associated with any form of IBD or specifically driven by one of its subtypes. Out of the 305 species tested for differential abundance, 208 exhibited a decrease in abundance in IBD or one of its subtypes, while 44 exhibited increased abundance (Q_FDR,Meta_ < 0.05). Most species were associated with IBD in general (n_up = 17, n_down = 148; Figure 1H, Supp. Figure 3). We found elevated levels of *Ruminococcus B gnavus*, *Flavonifractor plautii*, *Bacteroides fragilis* (and its closely related species), and *Clostridium Q symbiosum*, and a decreased abundance in multiple *Gemmiger* and *Alistipes* (A) species (Figure 1H, Supp. Figure 3).

Crohn’s Disease (CD) was found to be specifically associated with several species with strong abundance shifts compared to healthy control. We detected five *Enterocloster* species among the top 10 differentially abundant taxa that have elevated levels in CD, including *E. clostridioformis* and *E. boltea* (Figure 1H). As in previous reports (Chiodini et al., 2015; IBDMDB Investigators et al., 2019), we also found elevated levels of Pseudomonadota (previously named Proteobacteria) taxa, such as *Escherichia coli* and *Parasutterella* species. Several taxa showed substantial decreases in abundance specifically in CD, such as *Faecalibacterium* and *Blautia (A)* species, which is consistent with findings from previous amplicon-based analyses (Ma et al., 2022). Signals specific to ulcerative Colitis (UC) were less pronounced, with only six species showing a decrease in abundance, including four *Bacteroides* species. The taxa most prominently increased in UC included two *Acutalibacter* and three *Lachnospira* species (Figure 1H).

Looking into the differential abundances of 818 functional pathways (Figure 1I), we observed robust signals associated with CD, with 33 decreased pathways, and 259 increased pathways (Q_FDR,Meta_ < 0.05), compared to controls (Supp. Figure 3). The most strongly reduced pathway is the bile-acid 7alpha-dehydroxylation pathway (bai-pathway), responsible for the formation of LCA and DCA from chenodexycholate and cholate, respectively—a connection previously suggested in relation to CD (Larabi et al., 2023). We further identified a reduced abundance in the central fermentation pathway (CENTFERM-PWY), indicating a diminished production of butyrate, confirming past reports (Laserna-Mendieta et al., 2018). The elevated abundance of autoinducer AI-2 degradation suggests a potential shift towards a more selfish behavior in the microbiome, as autoinducers play a crucial role in quorum sensing within bacterial communities. Interestingly, we found that UC is associated with distinct bile acid metabolism pathways compared to CD, as the bile acid epimerization and iso-bile acid biosynthesis were significantly increased in abundance in UC compared to controls. This implies that variations in microbial bile acid metabolism may contribute significantly to the differences observed between Inflammatory Bowel Disease (IBD) subtypes. Finally, we identified histamine biosynthesis as being among the most prominent signals universally increased in IBD. While the role of histamine in IBD and other intestinal diseases is a subject of debate (Jutel et al., 2006), studies in mice undergoing experimental colitis suggested a potential involvement through the activation of innate immunity (Wechsler et al., 2018).

### Healthy siblings and children of IBD patients exhibit microbiome signatures of disease

The IBD-FC cohort provides the unique opportunity to explore potential shifts in the microbiota of healthy individuals that have a family history of IBD. To do this, family controls were categorized into groups based on the predominant IBD type in their respective families (FC CD and FC UC). Alternatively, FC were divided based on individual risk, with siblings and children of IBD-affected individuals designated as “high risk” (FC High) and other relatives (parents, partners, higher-degree relatives) as “low risk” (FC Low). All seven IBD incident cases included in the dataset were among the family members classified as FC High. In addition, two of the incident cases were not diagnosed with the IBD subtype predominant in their respective families. Our analysis revealed significant differences in community composition (Figure 2A) and alpha diversity (Figure 2B) between FC High and FC Low categories (P_alpha_ = 0.002, P_beta_ = 0.015). However, no difference in these two diversity metrics was found between FC CD and FC UC groups (Figure 2A & 2B).

**Figure 2:**
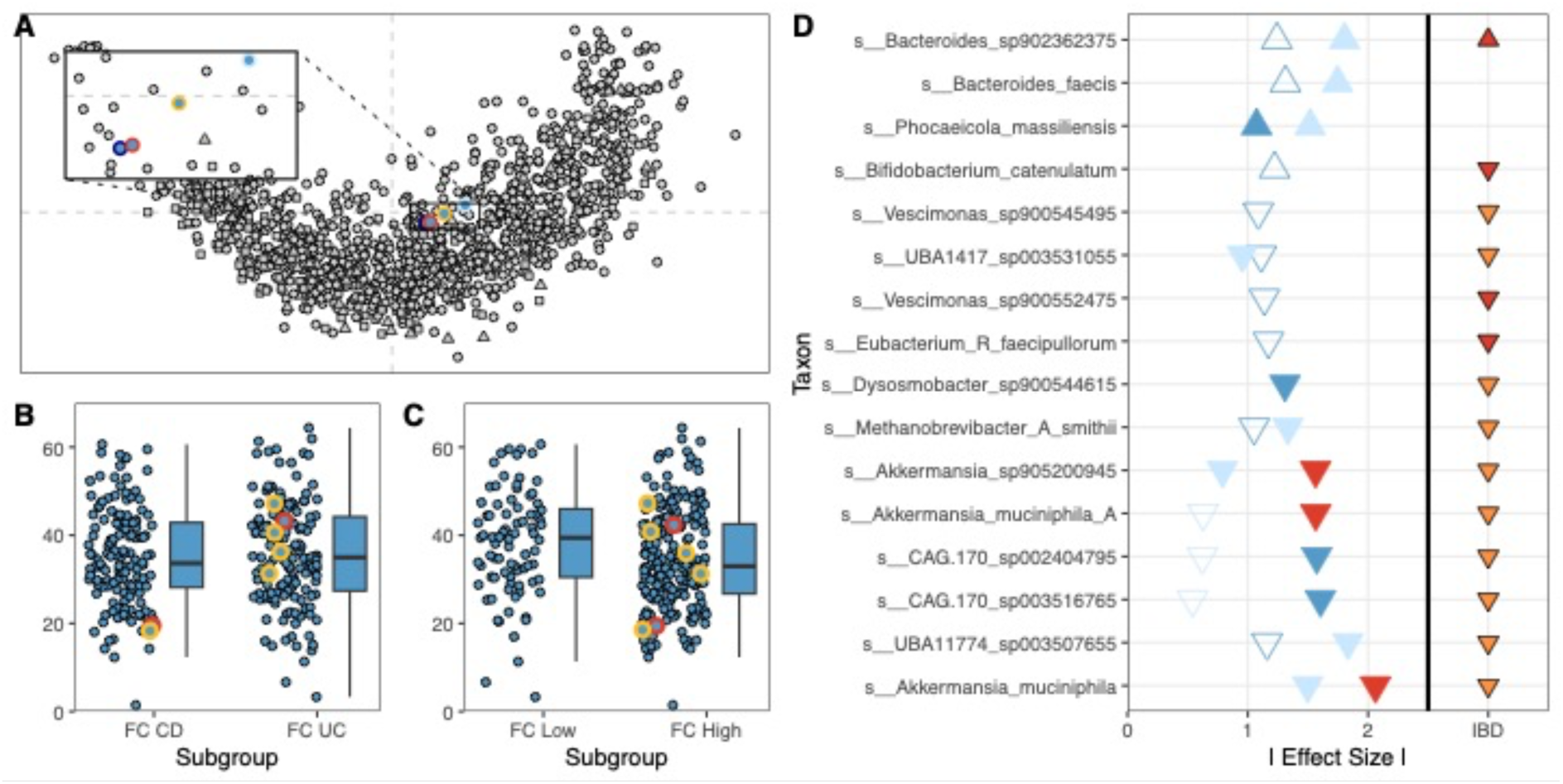
Family controls differ based on their risk to develop IBD, but not based on the IBD subtype predominant in their families. (A) Group centroids of healthy individuals from IBD-affected families (family controls = FC) grouped by IBD subtype predominant in the respective family (yellow = UC, red = CD; p_PERMANOVA_ > 0.05), and by disease risk (dark blue = high risk = siblings and children of IBD-affected individuals, light blue = low risk = parents, partners and higher degree relatives of IBD-affected individuals; p_PERMANOVA_ < 0.05). (B) Alpha diversity of FCs grouped by family-predominant IBD subtype (P > 0.05) and disease risk (P < 0.05). Pre-onset cases healthy at the timepoint of sampling are marked. (C) Differentially abundant species between low risk vs. high risk FCs (dark blue); between unrelated HC and high risk FCs (light blue); and between UC FCs vs CD FC (red). Filled shapes: q < 0.05, empty shapes: 0.05 < q < 0.1. The orientation of arrows reflects the effect direction of the difference in abundance with respect to the second group in the pairwise comparison (*e.g. A. muciniphila* is reduced in CD FCs compared to UC FCs, and reduced in FC High compared to HCs. The IBD column marks associations with IBD (orange) and IBD-subtypes (CD in red) as derived from the meta-analysis (Figure 1).

We also searched for microbial species that are differentially abundant in High FC vs. Low FC, in UC FC vs. CD FC, and in HC vs. High FC (Figure 2C), and compared these taxonomic hits against those found to have abundance signatures of IBD cases (Figure 1H). We found multiple taxa that are depleted in IBD or CD that are also depleted in either FC CD, such as *Akkermansia muciniphila*, or depleted in FC High compared to either FC Low or HCs, such as *Akkermansiav muciniphila* or *Methanobrevibacter A smithii* (Figure 2C). In addition, the CD-associated species *Bacteroides sp902362375* was found to be increased in FC High individuals compared to HCs. Altogether, these results suggest that FC High and CD FC individuals also exhibit microbiome signatures of IBD at the level of individual taxa or at the level of the community. These early shifts in the microbiome of FC individuals may potentially contribute to predisposing them to disease onset, in combination with other genetic and environmental factors, or serve as early markers for predicting disease risk. Further work will be needed to test and validate this hypothesis.

### Enterotypes and their dynamics associate with IBD subtypes

Microbiome community configurations, known as enterotypes, have been linked to various chronic inflammatory disorders affecting the gut and other organs (Valles-Colomer et al., 2019; Vieira-Silva et al., 2019). Here, we employed Dirichlet multinomial mixture (DMM) (Holmes et al., 2012) models to assess enterotypes based on genus-level abundances. We identified that a three-type model (ET1-3) explained best the diversity in community compositions in our data, based on Laplace approximation (Figure 3A). Next, we analyzed the distribution of our different disease and family control categories across the three enterotypes. ET1 was found to be the most prevalent among HC and FC Low individuals, particularly in the European cohorts, and poorly represented among CD individuals (Figure 3B). We observed a gradual loss of phylogenetic diversity from ET1 to ET3, ET2 being intermediate (Figure 3C). ET2 is more prevalent than ET1 among UC cases but is also more common in FC High compared to FC Low, again suggesting that FC High microbiomes are exhibiting signatures of IBD dysbiosis. Across all cohorts, ET3 is the most prevalent among CD cases, confirming their overall decreased alpha diversity (Figure 1). Examining community dynamics between timepoints in the German and USA cohorts, we observed that approximately two-thirds of individuals maintain their community configuration. Temporal shifts in enterotype most often involved transitions from or to the intermediate enterotype ET2, rarely occurring between ET1 and ET3 (Figure 3D). The high phylogenetic diversity of ET1 is associated with taxa that are usual markers of high phylogenetic diversity, including genera like *Prevotella* and *Methanobrevibacter A*. Many of ET1-associated taxa are also found to be depleted in IBD and IBD subtypes (Figure 3E). ET2 is associated with a few genera, specifically *Bacteroides*, suggesting that ET1 and ET2 represent the classical *Prevotella* and *Bacteroides* enterotypes, respectively, as found in previous studies (Costea et al., 2018). In contrast, ET3 represents a more ‘dysbiotic’ community state, characterized by high abundances of Escherichia coli, in addition to a low phylogenetic diversity.

**Figure 3:**
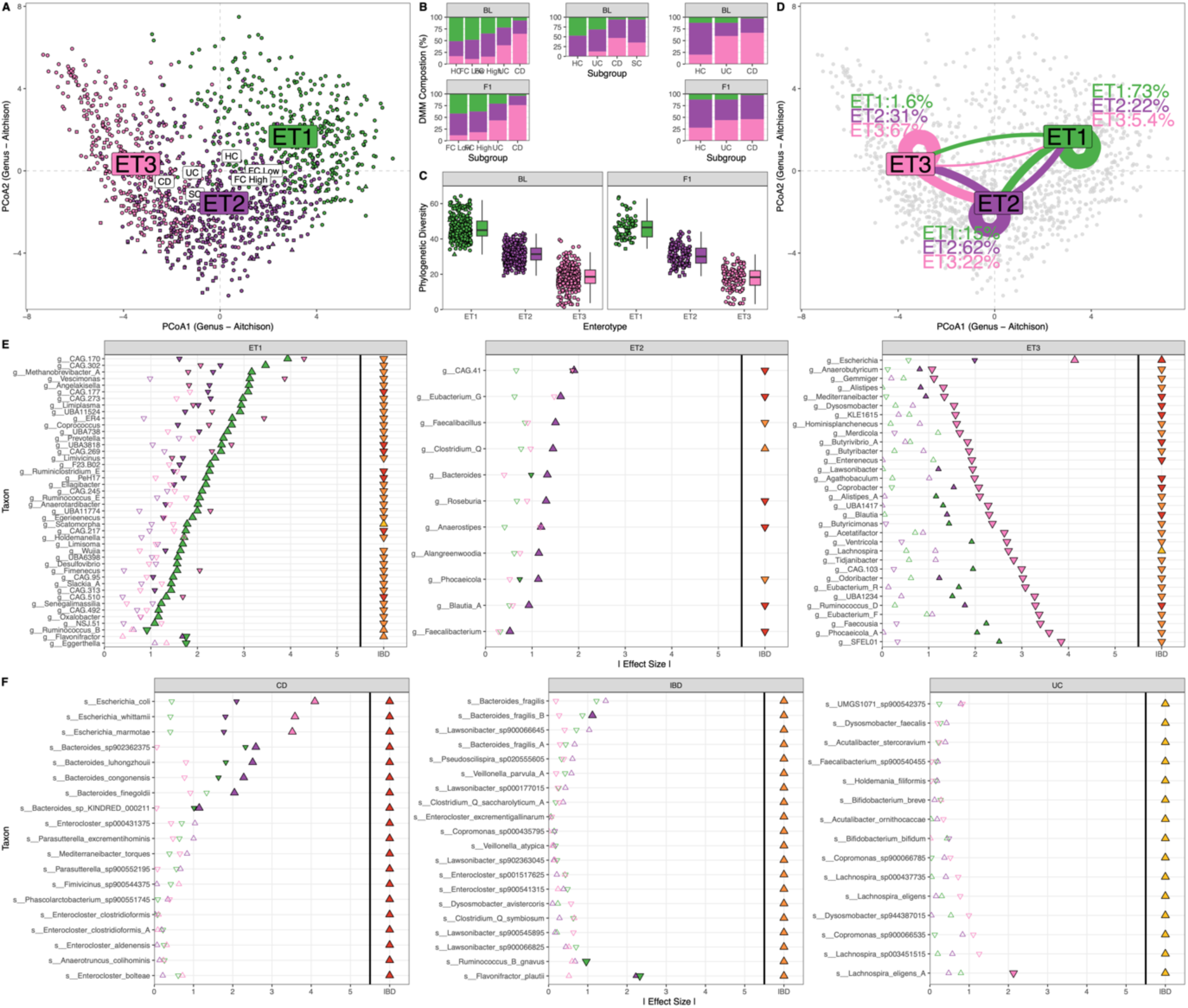
IBD is reflected in microbial community composition and diversity. (A) Distribution of derived enterotypes (ET) in the ordination plot. Colors throughout all panels – ET1: green; ET2: purple; ET3: pink. (B) Distribution of enterotypes within countries, study subgroups and timepoints. (C) Phylogenetic diversity by enterotype and timepoints across all cohorts. (D) Enterotype dynamics between timepoints. (E) Genus-level associations with the three enterotypes. Triangle symbols pointing downward or upward reflect decreased or increased abundances, respectively. Filled shapes represent significant adjusted P-values < 0.05. (F) Species enriched in IBD and IBD subtypes (CD on the left, IBD in the middle, and UC on the right), and their association with enterotypes.

While enterotypes are traditionally reconstructed based on genus-level profiles, as initially introduced using 16S rRNA gene amplicon data, metagenomic datasets have the potential to significantly enhance the resolution of enterotype analyses. We investigated the link between enterotypes and the 54 species positively associated with IBD (Figure 3F). Interestingly, species found increased in IBD or IBD subtypes are not all associated with specific enterotypes. However, three *Escherichia* species that are markers of CD are significantly enriched in the dysbiotic ET3 enterotype. Five CD-associated *Bacteroides* species were found enriched in the ET2 enterotype, with four of these being less abundant in ET1. None of the *Enterocloster* species that are enriched in CD (Figure 1F and G) are associated to a specific enterotype. As CD cases can be found across all enterotypes, this result suggests that CD-specific *Enterocloster* species have the potential to be used as microbial markers of CD with high sensitivity, independently of other characteristics of dysbiosis, such as the increase in *Escherichia* species. We did not find strong associations between UC microbial markers and enterotypes, with the exception of *Lachnospira eligens A,* which is significantly decreased in ET3. For taxa associated to IBD irrespective of disease subtypes, two taxa – *Bacteroides fragilis B* and *Flavonifractor plautii* – are found enriched in ET2. In addition, *Ruminococcus B gnavus* and *Flavonifractor plautii* are found in lower abundance in ET1. This analysis on individual taxa complements observations made at the community level and further suggests that ET2 represents an intermediary, and possibly predisposing, community state for IBD, compared to ET1.

### Decreased amino acid auxotrophies in Crohn’s disease associated communities

Next, we investigated the association between IBD and species-level or community-level amino acid (AA) auxotrophies. AA auxotrophies in microbial species and communities can be a marker of metabolic cross-feeding and their level in the gut microbiome were found to be associated with long term microbiome stability (Starke et al., 2023). We first measured species-level AA auxotrophies from genome-scale metabolic models that we built from our MAG collection (see Methods). We found strong signals of AA auxotrophy depletion in IBD in general and in CD in particular (Figure 4A). The lack of significant associations between auxotrophy and UC suggests that the significant depletion hits in IBD in general are mainly driven by the depletions found in CD. While there is a global trend for reduced auxotrophy in the microbiome of disease cases, we found that taxa that are auxotroph for glycine (Gly) are increased in abundance in IBD (Figure 4C). We next analyzed the abundance weighted mean number of auxotrophies per metagenomic sample in health and disease as a proxy for community-level measurement of auxotrophy. We found a significant reduction in auxotrophy levels in both UC and CD, with a stronger effect in CD individuals (Figure 4B). AAs with auxotrophy that are the strongest markers of disease are tyrosine (Tyr) auxotrophy (Figure 4D), but also tryptophan (Trp), which was significantly reduced in CD. Tryptophan metabolism was previously shown to be altered in (active) Crohn’s disease and was also found to be changed in abundance in the pathway-level analysis (Figure 1I). Community-level auxotrophy estimates are negatively correlated with phylogenetic diversity, independent of cohort subgroups (Figure 4F and 4G). Next, we wondered whether the loss of auxotrophy measured in disease was solely due to the loss of diversity. For this, we tested whether the number of AA auxotrophies in individual species is correlated with their association with IBD and IBD subtypes measured in the differential abundance analysis (Figure 1H). We found significant negative correlations between the number of auxotrophies in species and the effect size of for their association with IBD in general (P = 0.025) and with CD (P = 0.026) (Figure 4H). These results suggest that the observed reduced auxotrophies in disease are not simply an effect of depleted community diversity but are also driven by the increase in abundance of IBD and CD-associated taxa that have low number of auxotrophies. Overall, these results reveal shifts in the symbiotic relationships between gut bacteria in IBD and suggest that transitions towards more selfish metabolic and ecological behaviors occur in IBD microbial communities. Experimental approaches will be needed to validate this interpretation.

**Figure 4:**
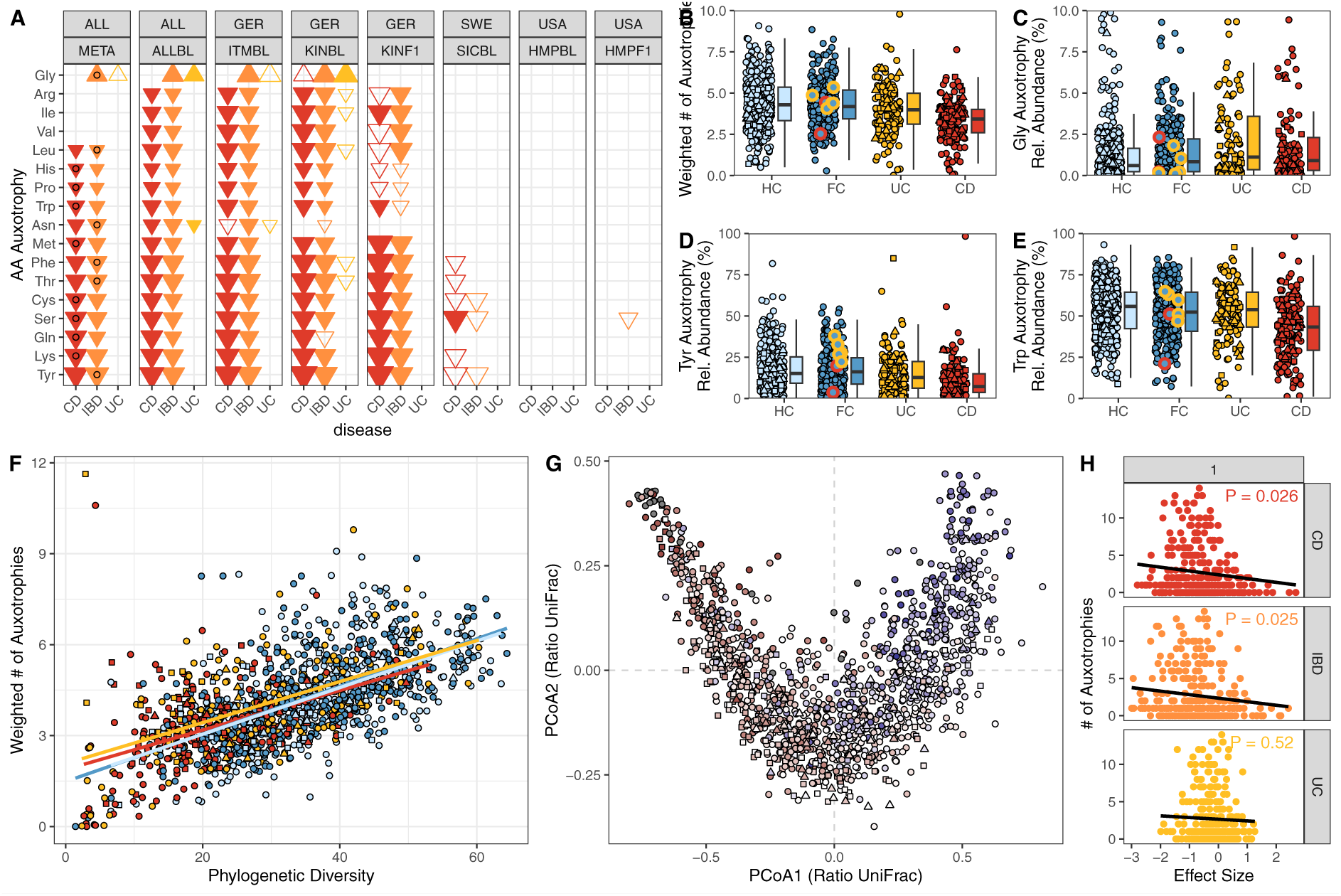
Widespread reduction of amino acid (AA) auxotrophies in IBD and specifically CD. (A) Results of the meta-analysis and individual cohort-level analyses for AA auxotrophies in IBD. Filled shapes represent adjusted P-values < 0.05, empty shapes unadjusted P-values < 0.05. Circles mark the association with the highest likelihood. (B). Abundance weighted levels of AA auxotrophies across study groups. Examples of increased (C: Gly) and reduced (D: Tyr; E: Trp) cumulative abundances of AA-auxotrophic clades in disease subgroups. (F) Correlation of abundance weighted auxotrophy levels and phylogenetic diversity per sample and by study group. (G) Distribution of abundance weighted auxotrophy levels across community compositions. (H) Correlation of species-level number of auxotrophies with effect sizes measured in the differential abundance analysis presented in Figure 1H.

### Longitudinal microbiome dynamics in IBD are changed at multiple scales

Longitudinal dynamics at the community level have been demonstrated to undergo alterations in IBD (IBDMDB Investigators et al., 2019). In our analysis of presence/absence data in the German cohort over approximately a 2-year span, we identified a significantly larger turnover of species at the species level in both IBD subtypes (Figure 5A). At the strain-level resolution, we observed that occurrences of strain replacements within species (popANI > 0.99999) were closely associated with the total number of shared species between timepoints (Figure 5B). However, when normalized by the total number of shared species between timepoints, we noted an increased relative population replacement in high-risk Family Controls (FCs) and Crohn’s Disease (CD) compared to low risk FCs (P_Bonferroni_ < 0.05; Figure 5C). Additionally, relative population replacements in Ulcerative Colitis (UC) compared to low risk FCs and CD compared to high risk FCs were also statistically significant (P_unadjusted_ < 0.05). These findings validate the existence of altered ecological dynamics specifically in Crohn’s disease at various taxonomic resolutions. Examining strain-level population replacement rates within individual species, we identified several *Bacteroides* species as well as *Agathobacter rectalis* to be differentially affected (P_Fisher_ < 0.05; Figure 5D).

**Figure 5:**
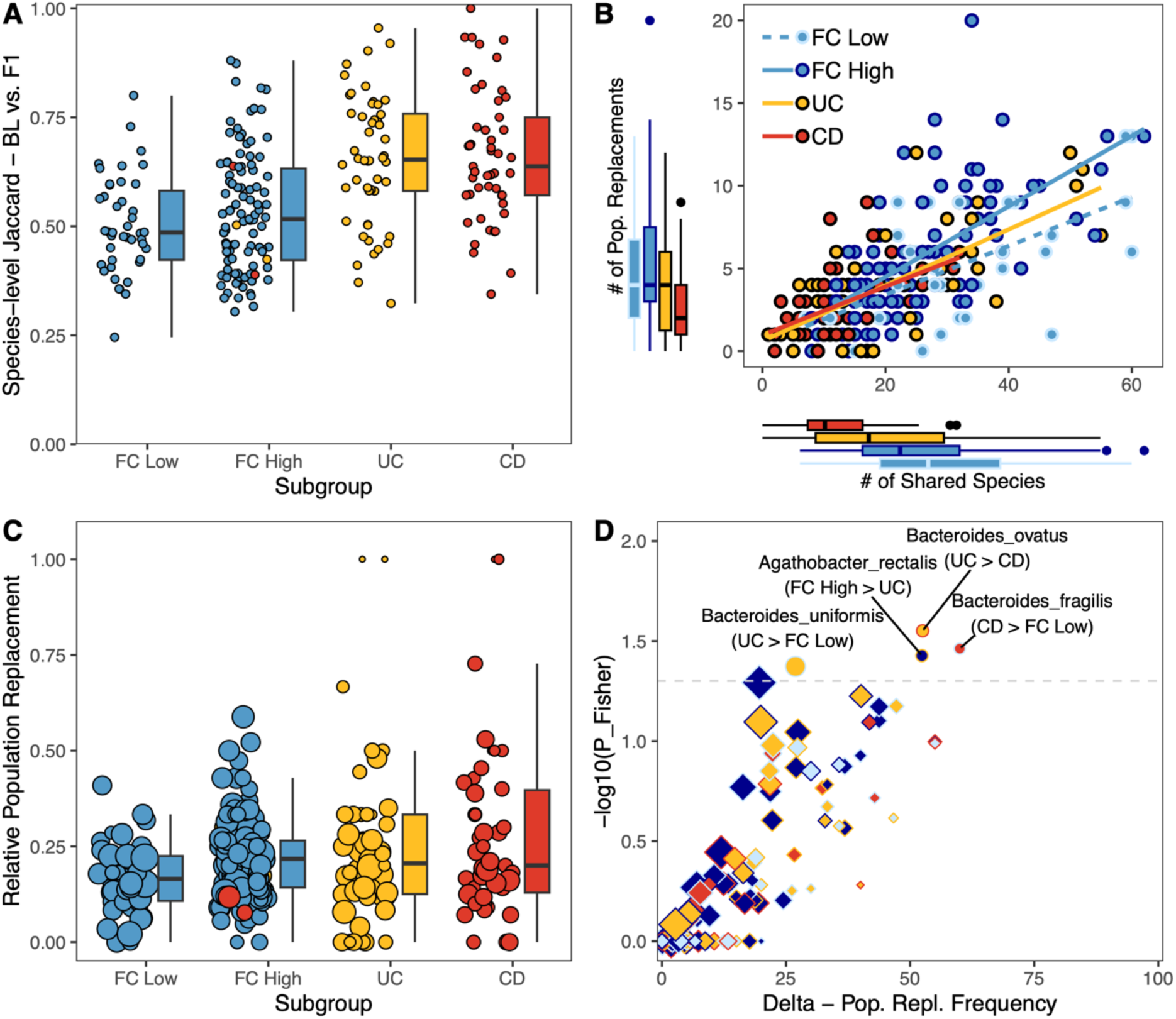
Altered ecological dynamics of individual gut bacteria in IBD subgroups. (A) Species-level jaccard distance between timepoints in the German cohort (B) Correlation of the number of shared species between timepoints and the number of strain-level population replacements derived from the inStrain analysis (C) Relative strain-level population replacement by study subgroup. (D) Pairwise comparison of population replacement frequencies within species and between study subgroups. Fill colors represent the subgroup for which the increased replacement frequencies were found, color of the outline represents the subgroup it was compared to. Shape sizes represent the total number of individuals the species was found shared across both timepoints.

## Discussion

We analyzed fecal metagenomic samples from 987 healthy controls and IBD patients from Germany, Sweden and the USA, including treatment-naïve cases and family controls, using meta-analysis approaches. We identified consistent cross-cohort community- and species-level associations at the level of taxa, pathways and auxotrophic metabolisms that are likely universal microbial markers of IBD and IBD subtypes. We also found microbial association patterns with IBD that reflect the geographic origin of the cohorts. Collectively, our findings suggest a decline in metabolic interactions in the IBD microbiome, specifically in CD-associated microbial communities, with an increased tendency toward self-sustained survival in an inflamed gastrointestinal tract. Whether this loss of altruism precedes dysbiosis or is its eventual outcome remains unclear. The building of the IBD-FC cohort aimed specifically at identifying early microbiome signatures of IBD. While the number of incident cases in our cohort remains too low to glean insights into these pre- and peri-onset microbiome dynamics, the inclusion and analysis of the microbiome of at-risk family controls revealed significant IBD-like shifts in the microbiome.

In our analyses, we combined likelihood-based prioritization of linear regression estimates to assign taxonomic associations to IBD subtypes, differentiating CD or UC-specific signatures from responses of microbes to IBD contexts shared between the two diseases, such as intestinal inflammation. Our analyses identified multiple *Enterocloster* species, including *E. clostridioformis* and *E. bolteae*, that are specifically enriched in CD and that may represent microbial markers of this IBD subtype. *Enterocloster* taxa have been somewhat overlooked in discussions about CD, potentially stemming from the limitations in resolution inherent in amplicon-based studies. In addition, previous investigations reported decreases in unclassified Lachnospiraceae (Ma et al., 2022), a family to which *Enterocloster* belongs, possibly contributing to the oversight of *Enterocloster* taxa, which then behaves opposite to the other clades within this family. Furthermore, their relatively recent re-classification from the (*Lachno*-)*Clostridium* genus into the *Enterocloster* genus may have added to this neglect. *Enterocloster* was identified and found associated to IBD in metagenomic data of a large Dutch cohort (Vich Vila et al., 2018), and to exhibit enriched transcriptional activity in IBD (IBDMDB Investigators et al., 2019). Yet, a clear association of specific *Enterocloster* species to CD was not investigated. Our high-resolution taxonomic calling approach that is based on dataset-specific MAGs found clear distinctions between different *Enterocloster* species, identifying some that are specifically associated with CD, such as *E. clostridioformis* and *E. bolteae.* Other *Enterocloster* species, such as *E. excrementigallinarium,* were found to be associated with IBD in general. The functional implications of CD-specific *Enterocloster* taxa in gut homeostasis or inflammatory processes will require dedicated experimental studies. Whether *Enterocloster* species, classified as obligate anaerobes, can cope with oxygenic environments in the inflamed gut also remains to be investigated in future work.

We also found multiple pathways specifically associated with CD. Among those, the elevated level of the degradation of autoinducer AI-2 pathway is of particular interest. AI-2 plays a crucial role in microbial quorum sensing and cell-to-cell communication (Pereira et al., 2013). Moreover, the intestinal epithelium secretes AI-2 mimics upon contact with bacteria, underscoring its involvement in cross-kingdom communication and the interaction between host and microbes (Ismail et al., 2016). Together with the observed decrease in AA auxotrophies in CD, these findings reinforce the idea of an inflammation-associated shift of the microbiome toward a more ‘selfish’ and less cooperative microbial community.

The microbiome of individuals affected by UC exhibits less pronounced differences compared to healthy controls than those observed in CD. Despite the general reduction in diversity associated with IBD in general and widespread taxonomic changes also applicable to UC, subtype-specific alterations are less prominent. Only six species are specifically depleted in UC, including four within the genus *Bacteroides*. This finding starkly contrasts with the positive association of *Bacteroides fragilis* and other *Bacteroides* species with IBD in general (Figure 1I), highlighting the significance of species-level resolution in microbiome studies. This level of granularity enables the investigation of distinct and opposing changes within a single microbial genus associated with the disease. Nonetheless, the UC-specific increases in species belonging to the genera *Acutalibacter*, *Copromonas* (both Oscillospirales), *Lachnospira*, *Dysosmobacter* (both Lachnospiraceae), and *Bifidobacterium* (*B. breve* and *B. bifidum*) could potentially elucidate UC-related processes and contribute to the microbiome-based differentiation of IBD subtypes. The notable rise in *Bifidobacterium breve* is particularly intriguing, presenting a significant departure from previous reports that identified *B. breve*, along with other *Bifidobacterium* species, for its potential to ameliorate DSS-induced colitis in mice (Singh et al., 2020). However, additional studies have documented strain-specific effects for *B. breve*, and human probiotic intervention trials have indicated no discernible impact of oral *B. breve* supplementation on relapse-free survival (Matsuoka et al., 2018). None of the other identified clades have been identified as associated with UC in any metagenome-based analysis, and they are generally insufficiently characterized in the context of the gut microbiome. Their role in UC awaits further clarification.

Pathways exhibiting increased abundances in UC hint at altered bile acid metabolism, specifically linked to shifts in microbial communities. Previous studies have demonstrated modified compositions of primary and secondary bile acids in individuals with IBD, particularly during episodes of acute colitis (Thomas et al., 2022). We found that bile acid epimerization and iso-bile acid biosynthesis pathways are increased in UC. Iso-bile acids are less toxic to bacteria (Devlin & Fischbach, 2015), their stable 7-oxo-intermediates are thought to actively affect host steroid hormone levels (Odermatt & Klusonova, 2015) and are non-active agonists of the farnesoid X receptor (FXR), blocking downstream signaling and production of antimicrobial peptides, such as cathelicidin (Campbell et al., 2012). Thus, bile acid epimerization can manipulate host defense mechanisms in multiple ways and may contribute to pathogenesis in UC (Ridlon et al., 2016).

Regardless of the subtype, the microbiome of IBD cases exhibited a reduced abundance of over 150 microbial species compared to healthy controls. Other species, such as *Ruminococcus B gnavus, Flavonifractor plautii, Clostridium Q symbiosum,* and *Bacteroides fragilis* including two closely related strains *B. fragilis A* and *B* were found in increased abundances in the samples of IBD-affected individuals. These taxa have previously been identified to be associated with IBD (Hassouneh et al., 2021; Pisani et al., n.d.). *R. gnavus* has been extensively reported to be associated with multiple intestinal and extra-intestinal disorders (Crost et al., 2023) and was shown to actively promote inflammation through TLR4 dependent TNF-alpha secretion in response to the polysaccharide glucorhamnan (Henke et al., 2019). *F. plautii* and *C. symbiosum* were implicated in bloodstream infections (Elsayed & Zhang, 2004; Karpat et al., 2021), and *Bacteroides fragilis* is known to be an opportunistic pathogen (Wexler, 2007). *B. fragilis* virulence is likely expressed in response to oxygen exposure via the oxyR regulon (Rocha et al., 2003; Sund et al., 2008). The increased abundance of ROS in IBD-associated intestinal inflammation is potentially a trigger for the switch of *B. fragilis* from a commensal to pathogenic lifestyle. Yet, the contribution of *B. fragilis* to IBD progression remains to be experimentally demonstrated.

Intriguingly, we found that the hyaluronan degradation pathway (PWY-7645) was among the most increased in IBD. Hyaluronan is known to be deposited in the extracellular matrix (ECM) at inflamed sites, and is actively involved in the recruitment of immune cells (Petrey & de la Motte, 2019). It was also previously shown that IBD-associated bacteria are able to degrade ECM components, including hyaluronan (Porras et al., 2022). How these dynamics influence IBD onset and progression needs to be investigated. Yet, the dysregulation of hyaluronan deposition and catabolism were shown to be involved in inflammatory processes and coagulation, increasing the risk for microvascular occlusion and thrombosis, which are known comorbidities in IBD (Petrey & de la Motte, 2019). Additionally, our analyses could confirm many associations previously reported in the microbiome of IBD patients, such as the mucus-degrading *Akermansia muciniphila, Alistipes spp.,* the *Prevotella copri* complex*, Gemmiger spp.,* and *Methanobrevibacter A smithii* (Vich Vila et al., 2018). The methanoarchaeon *M. smithii* is a known marker of healthy anaerobic gut microbiomes, as demonstrated by several studies focusing on the evolution of mammalian gut microbiomes (Rühlemann et al., 2023). By producing methane as a metabolic product in anoxic conditions through the fermentation of bacterial primary products, such as acetate, methanol, hydrogen and carbon dioxide (Thauer, 1998; Thauer & Shima, 2006), it can engage in syntrophic interactions with a broad range of bacteria (Samuel et al., 2007; Samuel & Gordon, 2006). Several studies demonstrated more efficient polysaccharide fermentation in the human intestine methanogenesis through the presence of *M. smithii*, as it consumes various bacterial fermentation products thereby lowering the overall redox potential (Miller et al., 1984; Samuel et al., 2007; Samuel & Gordon, 2006). Methane production is also linked to low transit time and anti-inflammatory effects in the colon (Boros et al., 2012; Pimentel et al., 2006). Finally, reduced *M. smithii* abundances are inversely correlated with the abundance of sulfate-reducing bacteria (Ghavami et al., 2018). These microorganisms produce toxic hydrogen sulfide, which was also implicated in IBD pathogenesis (Ghavami et al., 2018). Owing to the multiple key metabolic and physiological functions that *M. smithii* perform in the gut ecosystem, it was previously proposed that it may serve as a biomarker for IBD (Ghavami et al., 2018). Our study, which revealed reduced abundances of *M. smithii* in high-risk FCs compared to low-risk FCs or HCs provide additional support to this idea, as it suggests that the loss of *M. smithii* defines a pre-dysbiotic state that is on the path to IBD dysbiosis.

Our study emphasizes the potential to make novel discoveries into the role of the microbiome in IBD by employing meta-analysis approaches on metagenomic datasets derived from both prospective and family cohorts. The future expansion of the IBD-FC cohort is poised to play a pivotal role in enhancing our comprehension of the intricate interactions among hosts, the environment, and the microbiome in the pathogenesis of IBD.

## Methods

### Cohort description and sampling

The IBD-FC is a German nationwide, prospective cohort study collecting both, questionnaire data and biomaterial from IBD patients and their (affected and unaffected) family members. The recruitment of study participants started in October 2013 and is still ongoing. Follow-up information and new biomaterial samples are collected prospectively at intervals of approximately two years. All participants, including the IBD patients, provided questionnaire data and biomaterial (blood, stool, hair) at baseline and after every 2 years of follow-up. Samples were taken at home by the participants and sent by mail to the Microbiome Laboratory of the Institute of Clinical Molecular Biology for further analysis. In addition, physician-administered questionnaires have been collected from IBD patients and onset cases). Every study participant has given written informed consent on forms that are age-adapted. For participants under the age of 18 years, the informed consent must also be signed by their parents. IBD-FC IBD diagnoses were curated to be grouped into the correct subtype based on questionnaire and physician data of all available timepoints, using the latest diagnosis. The study was conducted in accordance with the Declaration of Helsinki. Ethics committee of the Medical Faculty of Kiel University gave ethical approval for this work (AZ A117/13).

Subjects from Northern Germany from a blood donor cohort were recruited by mail and asked to provide stool samples and detailed food, lifestyle and medical history questionnaires. Samples were taken at home and sent by mail within 24 h after sample collection to the Microbiome Laboratory for further analysis. After arrival stool samples were aliquoted and stored at −80 °C until further processing. The study was conducted in accordance with the Declaration of Helsinki. Ethics committee of the Medical Faculty of Kiel University gave ethical approval for this work (AZ A103/14). Metadata were assessed for the presence of chronic diseases and samples were included in case of missing questionnaire data, and if individuals were affected by IBD, irritable bowel syndrome, primary sclerosing cholangitis, primary biliary cirrhosis, or any type of diabetes. In total 112 out of 456 samples were excluded from the ITM dataset.

The Swedish Inception Cohort in IBD (SIC IBD) was previously described and includes treatment-naïve patients, between 20–77 years of age (Bergemalm et al., 2021).

Quality-filtered metagenomic data from the human microbiome project (HMP) were downloaded for a subset of individuals for which a baseline sample and a sample at 6 months (180 day +/-30 days) with > 3 x 10∧6 reads were available based on the information provided by the curatedMetagenomicData package for R (Pasolli et al., 2017), study_name == “HMP_2019_ibdmdb”.

Data and material of the IBD-FC cohort are available upon the submission of a research proposal to https://portal.popgen.de/.

Data of the ITM cohort will be uploaded to ENA upon acceptance of this manuscript at a peer-reviewed journal.

### Metagenome sequencing and data processing

#### Sample preparation and sequencing

Total fecal DNA samples were extracted from 200mg stool aliquots starting with bead beating in a SpeedMill PLUS (Analytik Jena AG) for 45 s at 50 Hz mixed with 1.1 mL ASL buffer in 0.70mm Garnet Bead tubes (Qiagen). Subsequently, samples were heated to 95°C (5 min) followed by centrifugation. 200 µl of the resulting supernatant were used for automated DNA extraction with the QIAcube system (Qiagen) using the QIAmp DNA Stool Mini Kit (Qiagen). DNA quality was assessed by Qubit and Genomic DNA ScreenTape (Agilent). Illumina Nextera DNA Library Preparation Kit was used to construct shotgun metagenomic libraries, and subsequently sequenced with 2 × 150 bp reads on a NovaSeq 6000 system (Illumina).

#### Quality control and binning

All metagenomic data were processed using the TOFU-MAaPO pipeline for metagenomic data (https://github.com/ikmb/TOFU-MAaPO). Briefly, sequencing data were quality controlled to remove low quality reads, sequencing adaptors and PhiX spike-in reads using BBTools (Bushnell, 2014). Human host-derived reads were removed using Bowtie2 (Langmead & Salzberg, 2012) and the hg19 version of the human reference genome. HMP data were already quality filtered and the above steps were skipped for these samples. Assembly of metagenomic contigs was performed with Megahit (D. Li et al., 2015) using k=27….127 for each samples individually, retaining contigs > 2kbp. Reads were mapped to the resulting contigs using minimap2, resulting SAM files were converted to ordered BAM files with samtools and per-contig coverages were calculated using the jgi_summarize_bam_contig_depths (H. Li et al., 2009). MaxBin2 (Wu et al., 2016), Metabat2 (Kang et al., 2019), and CONCOCT (Alneberg et al., 2014) were used for binning. Additionally, samples were allocated to groups of 37-54 based on their sample IDs for binning with VAMB (Nissen et al., 2021). Within each group, contigs were concatenated and reads were mapped to this catalog using minimap2, following the same procedure as described above to create VAMB input files. GTDBtk (Chaumeil et al., 2022) bac120 and ar53 marker genes were identified from the prodigal (Hyatt et al., 2010) output of the contigs in --meta mode, which were annotated using HMMer (Eddy, 2011) and used for bin scoring, refinement and estimation of completeness/contamination with the MAGScoT tool (Rühlemann et al., 2022), resulting MAGs with a score > 0.5 and contamination < 10% were retained for downstream analyses.

#### Species-level genome bin (SGB) catalog

We applied a multi-step clustering to the collection of MAGs to minimized SGB catalog inflation due to low-quality MAGs. Within groups, MAGs were dereplicated at 97% identity using dRep (Olm et al., 2017), for each cluster the highest scoring MAG was chosen as representative. Pairwise mash distances of good quality (scores >= 0.7) cluster representatives across all groups were calculated and used for primary single-linkage clustering at 90% similarity. Within primary clusters, Average nucleotide identities (ANI) were calculated using fastANI (Jain et al., 2018) and average linkage clustering was performed resulting in a set of high-quality SGBs selecting the highest scoring MAGs as SGB representatives . Medium quality MAGs (MQ MAGs; 0.5 < score < 0.7) were compared against SGB representatives using fastANI and MQ MAGs with >= 95% ANI to an SGB were assigned to its cluster. MQ MAGs without matching SGBs (all ANI < 0.95) were subjected to ANI-based average-linkage clustering as described above and added to the SGB catalog only if they were recovered from at least two samples across the dataset, removing singleton MQ MAGs from the dataset. SGB representatives were subjected for taxonomic annotation using GTDBtk (v2.1.0; (Chaumeil et al., 2022)) and GTDB release 214 as reference database. GTDBtk marker gene alignments of representative SGVs were used to reconstruct a phylogenetic tree using IQTREE (Minh et al., 2020; Nguyen et al., 2015). Representative SGB genomes were concatenated and indexed as reference for salmon (Patro et al., 2017).

#### Abundance estimation

Clean metagenomic reads were mapped against the SGB catalog using salmon quant in --meta mode and using the –validateMappings option (Patro et al., 2017). Resulting contig-level mappings were used to estimate SGB-level abundances. For this, a contig was considered to be present in a sample if it had at least 10% coverage, calculated as the number of mapping reads x 2 x 150bp / effective contig length. An SGB was considered present if at least 20% of the cumulative effective contig lengths qualified as present and at least 1000 reads and 250 counts per million (CPM) were mapped to the SGB. Using GTDBtk annotations of SGBs, domain- to species-level abundances were calculated as cumulative CPM abundances of the respective level and clade. CPMs are genome-size and sequencing-depth adjusted with an upper bound of 10∧6 per sample, thus representing relative abundances. At each taxonomic level, CPMs were used to calculate centered-log ratio (CLR) abundances to account for compositionality of microbiome date, applying the clr() function of the compositions package for R and adding a pseudocount of 1 (van den Boogaart & Tolosana-Delgado, 2008). Resulting negative CLR values were set to 0.

#### Gene/protein catalog

Prodigal (Hyatt et al., 2010) outputs of predicted amino acid sequences of coding regions for all samples were concatenated and clustered at 95% identity using mmseqs v2 in “easy-linclust” mode with options “--cov-mode 1 -c 0.8 --kmer-per-seq 80”. Resulting representative sequences were renamed to carry standardized IDs and subjected to annotation with emapper.py (Cantalapiedra et al., 2021) using the eggNOG 5.0 (release 220425; (Huerta-Cepas et al., 2019)) database as reference. Further, the 95% identity catalog was used to create catalogs at coarser-level resolution of 80% and 50% identity. EggNOG annotations of 95% identity clusters were collapsed to represent broader-level protein clusters. The 50% identity catalog was used to create a DIAMOND reference (Buchfink et al., 2015) to which metagenomic reads were mapped and gene/protein-level functional CPM abundances were estimated.

#### Metabolic modelling and prediction of amino acid auxotrophies

Amino acid auxotrophies were predicted using computational metabolic modelling based on genome-scale metabolic networks as described previously (Starke et al., 2023). In brief, genome-scale metabolic networks were reconstructed for species-representative MAGs using *gapseq* version 1.2 (Zimmermann et al., 2021). Amino acid auxotrophies were predicted using flux balance analysis, where the objective function was defined as the flux through the biomass formation reaction. An organism was defined as auxotrophic for a specific amino acid if the corresponding metabolic model could form biomass in the original medium but not in the medium without the amino acid of interest. Flux balance analysis was performed in R (v4.3.1), the R package *sybil* v2.2.0 (Gelius-Dietrich et al., 2013), and the IBM ILOG CPLEX optimiser as linear programming solver.

#### Quantifying the abundance of predicted metabolic pathways

Quantitative compositional pathway abundances per metagenome sample were estimated by combining TPM values for species-representative MAGs and the MAGs’ pathway presence/absence predictions from the metabolic modelling approach (see above). Specifically, the abundance *q* of pathway *i* in sample j was calculated as

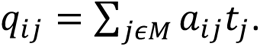

*M* is the set of all MAGs, *a*_ij_ is the prediction of pathway *i* in organism *j* (*0* if absent; *1* if present), and *t_j_* is the TPM of MAG *j*.

### Downstream data processing and statistical analysis

All further processing and statistical analyses were carried out in R 4.2.1 (R Core Team, 2022) and utilitzing the tidyverse collection of packages (Wickham et al., 2019). Alpha diversity was assessed as Faith’s phylogenetic diversity using SGB-level abundances and the reconstructed phylogenetic tree in the pd() function of the picante package (Kembel et al., 2010). Differences in alpha diversity between groups were assessed using linear regression. Ordinations are based on ratio UniFrac distances of SGB-level abundances and the reconstructed phylogenetic tree (Lozupone et al., 2011; Wong et al., 2016). Differences in beta diversity were assessed using permuatitional multivariate analysis of variance using distances matrices implemented in the adonis2() function of the vegan package (Oksanen et al., 2022).

### Differential abundance analysis

Stratified by country, abundance differences of taxonomic groups in healthy controls and IBD-affected individuals were assessed using CLR-transformed abundances and linear models with age, sex and BMI as covariates, except for the SIC-IBD cohort where BMI data were not available. Tested were all taxa with overall prevalence of >= 30% or within-group prevalence >= 20%. Group comparisons performed were (1) all IBD vs HC, (2) CD vs. all non-CD (HC + UC), and (3) UC vs. all non-UC (HC + CD). Similarly, assessing differences in family controls and IBD-affected individuals, linear mixed models were used with ‘family ID’ as random effects variable in addition to the fixed effects named above. For the cross-country comparison of HC and IBD cases, linear mixed models were used with ‘country’ as random effects variable and age + sex as fixed effect variables. This model was used to sort taxa into being enriched/depleted with either IBD in general or one of its subtypes based on the log-likelihood of the model. Inverse-variance-weighted meta-analysis was used on the analyses of HC and IBD-affected individuals to obtain final association statistics. P-values from the meta-analysis and individual linear models were FDR- or Bonferroni-corrected for multiple testing, respectively. Pathway abundance-based analyses were performed as described above for taxonomic groups, however abundances were log-transformed as these data are not compositional and CLR-transformation would not be appropriate.

### Enterotype inference and analysis

The number of enterotypes was assessed based on HCs from all countries using genus-level abundances and Dirichlet multinomial mixtures implemented in the dmn() function of the DirichletMultinomial package for R (Holmes et al., 2012). Optimal number of clusters between 1 and 8 were assessed using the Laplace estimator for goodness-of-fit. The respective model with k=3 clusters was used to classify non-HC samples into one of three enterotypes. Differential abundances of taxonomic groups between enterotypes were assessed using linear models, including age + sex + BMI as covariates and comparing samples from one enterotype against the other two. Assignments of enterotype-specific increase or decrease of abundances were based on a Bonferroni-corrected P-values < 0.05 and highest log-likelihood.

### Auxotrophy analysis

Differential abundance analysis of amino acid auxotrophic microbes is described above. Mean per-sample auxotrophies were estimated from SGB-level auxotrophy counts weighted by their abundances. Auxotrophy abundance-based analyses were performed as described above for taxonomic groups, however abundances were log-transformed as these data are not compositional and CLR-transformation would not be appropriate.

### Analysis of strain-level dynamics

MAGs of samples for which two timepoints were subjected to dereplication with dRep to obtain a set of non-redundant SGBs per sample, the SGB representative sequence was chosen based on its completeness and contamination and concatenated in a per-sample SGB (sampSGB) catalog. This catalog was used as bowtie2 reference, reads from both timepoints were mapped against the reference individually and the resulting SAM file was used as input in inStrain (Olm et al., 2021) in ‘profile’ mode with an additional file listing contig-to-sampSGB mappings. The resulting profiles were then compared in the inStrain ‘compare’ mode. An sampSGB was considered present at a timepoint if it had a higher than 3x coverage. A strain-level population replacement was present in the case of population average nucleotide identity (popANI) < 0.99999 between timepoints. Pairwise species-level enrichments of population replacements between groups were assessed using Fisher’s exact test based on the counts of species-sharing and detected population replacements. Species were included in the comparison for a subgroup if they were found shared between timepoints in at least 5 samples in the respective groups.

## Data Availability

Data and material of the IBD-FC cohort are available upon the submission of a research proposal to https://portal.popgen.de/.
Data of the ITM cohort will be uploaded to ENA upon acceptance of this manuscript at a peer-reviewed journal.

## Acknowledgements

We would like to thank all patients and probands involved in the sampling of biomaterials for this study. We are thankful to the staff of the IKMB microbiome laboratory for processing the stool samples. This work was supported by the DFG Research Infrastructure NGS_CC (project 407495230) as part of the Next Generation Sequencing Competence Network (project 423957469). NGS analyses were carried out at the Competence Centre for Genomic Analysis (Kiel).

